# Beneficial Association of HDL Cholesterol With Reperfusion Injury And Functional Outcome After Thrombectomy For Stroke

**DOI:** 10.1101/2024.10.14.24315500

**Authors:** Annahita Sedghi, Sonja Schreckenbauer, Daniel P. O. Kaiser, Ani Cuberi, Witold H. Polanski, Martin Arndt, Kristian Barlinn, Volker Puetz, Timo Siepmann

## Abstract

**Background:** Animal studies suggest that high-density lipoprotein cholesterol (HDL-C) protects against reperfusion injury. We hypothesised that higher serum HDL-C levels would have a protective effect against cerebral reperfusion injury in human stroke survivors treated with thrombectomy.

**Methods:** We included consecutive patients from our prospective anterior circulation large-vessel occlusion (acLVO) registry who underwent thrombectomy between January 2017 and January 2023 at a tertiary stroke centre in Germany in a propensity score-matched analysis. We assessed the association between serum HDL-C levels and imaging indices of post-interventional reperfusion injury according to the Heidelberg Bleeding Classification as well as 90-day functional outcome quantified by the modified Rankin Scale (mRS). We performed sensitivity analyses using multivariable lasso logistic and linear regression adjusted for demographic, clinical and imaging characteristics.

**Results:** Out of 1702 patients assessed for eligibility, 807 acLVO patients treated with thrombectomy (420 females, median age 77 years [66-84, IQR]) were included. Reperfusion injury reduced the probability of a favourable functional outcome (90-day mRS 0-2) by 14.8% (ß=0.15; 95% CI [0.06;0.24]; *p*=0.001. A serum HDL-C level above the median (1.15 mmol/L) decreased the probability of reperfusion injury by 13.6% (ß=-0.14; 95CI% [−0.22; −0.05]; *p*=0.002) and increased the probability of favourable functional outcome by 13.2% (ß=-0.13; 95CI% [-0.22;-0.05]; *p*=0.003). In sensitivity analyses, higher HDL-C levels were associated with lower odds of reperfusion injury (adjusted OR 0.62; 95% CI [0.43;0.88]; *p*=0.008) and emerged as a predictor of a favourable functional outcome (adjusted OR 0.60; 95% CI [0.40; 0.90]; *p*=0.015).

**Conclusions:** In patients undergoing thrombectomy for acLVO, higher serum levels of HDL-C were associated with a reduced probability of reperfusion injury and favourable functional outcome at 90 days.

## Introduction

Thrombectomy for anterior circulation large vessel occlusion (acLVO) has revolutionised stroke care ever since randomised clinical trials published in 2015 as well as their meta-analytic synthesis have confirmed the efficacy and safety of the treatment; an observation that has more recently been reproduced in an extended time window of up to 24 hours from stroke onset, large core infarcts and basilar artery occlusion.^1,2^ Observational research and sub-analyses of randomised trials have shown that the beneficial effects of thrombectomy may be attenuated by reperfusion injury.^3^ Abrupt reperfusion of ischaemic brain tissue can lead to local inflammation and oxidative stress, endothelial damage and oedema, and consequently to breakdown of the blood-brain barrier with the possible consequence of haemorrhage. This complication has been reported after carotid endarterectomy and stenting and, more recently, after thrombectomy.^4–6^ In brain tissue affected by reperfusion injury, haemorrhage may manifest as haemorrhagic transformation, parenchymal or subarachnoid haemorrhage.^7–9^ Radiologically confirmed haemorrhagic reperfusion injury occurs in up to one third of patients after thrombectomy with partial or complete reperfusion, worsening functional outcome.^7^ To date, the optimal strategy for preventing of this complication is unknown. There remains an urgent need for therapeutic targets to protect brain tissue from reperfusion injury after thrombectomy.

Animal studies have shown that high-density lipoprotein cholesterol (HDL-C)-based treatment exerts direct vasculoprotective and neuroprotective effects^10^, preserves the integrity of the blood-brain barrier after transient intraluminal occlusion of the middle cerebral artery^12^, and reduces the risk of bleeding after treatment with tissue plasminogen activator.^13^ To our knowledge, the effects of HDL-C on cerebral reperfusion injury after thrombectomy in humans are unknown. We tested the hypothesis that patients with higher serum HDL-C levels undergoing thrombectomy for acLVO would have a lower risk of haemorrhagic reperfusion injury than those with lower HDL-C levels.

## Methods

### Study Design and Patients

We included all adult acLVO patients from our prospective endovascular treatment registry who underwent thrombectomy at a tertiary stroke centre between 01/2017 and 01/2023 in a retrospective cohort study. This study report conforms to the Strengthening the Reporting of Observational Studies in Epidemiology (STROBE) statement.^14^ The *STROBE checklist* is provided in the *Supplementary Material.* We excluded patients with an incomplete or missing lipid panel, missing follow-up cranial imaging, unknown onset of symptoms or time of last seen well, unknown baseline National Institute of Health Stroke Scale (NIHSS) score or missing modified Rankin Scale (mRS) score at 90 days. Definitions of patient characteristics are provided in the *Supplementary Methods*.

### Prospective Thrombectomy Registry

Our registry of thrombectomy-eligible patients includes detailed data on demographic characteristics, cardiovascular risk profiles, premorbid conditions, chronic comorbidities, medications, stroke etiology as classified by Trial of Org 10172 in Acute Stroke Treatment (TOAST), modified treatment in cerebral infarction (mTICI) score, neurological deficits as quantified by the National Institute of Health Stroke Scale (NIHSS), and functional outcome as assessed by the modified Rankin Scale (mRS) at admission, discharge, and 90 days. Fasting serum lipid profiles were obtained during the stay on the stroke unit. Details of the registry are provided in *Supplementary Methods*, and the details of the collection of baseline and outcome parameters are provided in *Supplementary Table S1*, respectively.

### Assessment of Imaging and Clinical Outcomes

We considered reperfusion injury to be present if at least one of the key radiological features of reperfusion injury was evident on follow-up cranial computed tomography or cranial magnetic resonance imaging within 24 hours of thrombectomy. These key features included any haemorrhagic transformation, defined as categories HI1 and HI2 of the Heidelberg Bleeding Classification, and any parenchymal (PH1 and PH2) or subarachnoid (class 3c) haemorrhage involving the ischaemic brain region as previously proposed.^9,15^ All brain scans were evaluated by board-certified neuroradiologists. Successful recanalisation was defined as an mTICI score of 2b or higher. Functional outcome was assessed by telephone interview 90 days after thrombectomy. Favourable functional outcome was defined as a modified Rankin Scale (mRS) score of 0 to 2 and poor functional outcome as an mRS score ≥ 3 at the time of this follow-up. Neurological deficits were assessed on admission and discharge using the National Institutes of Health Stroke Scale (NIHSS).

### Statistical Analysis

Independent continuous variables were tested for normality using descriptive and analytical (Shapiro-Wilk test) criteria. Differences between groups in demographic, clinical, imaging and procedural characteristics were assessed using Fisher’s exact test for binary data and Kruskal-Wallis test for categorical or non-normally distributed continuous data, where appropriate.

For the main analysis, propensity score matching was carried out. Patients were divided into two groups based on their serum HDL-C level, with the cut-off value set at the median HDL-C level of the entire study population. Accordingly, low HDL-C was defined as an HDL-C level < 1.15 mmol/L and high HDL-C ≥ 1.15 mmol/L. Covariates were selected by multivariable logistic regression including clinically relevant covariates, i.e. age, sex, premorbid dependency, chronic disease that may affect functional independence, baseline NIHSS score, Alberta Stroke Program Early CT Score (ASPECTS), occlusion site, concomitant extracranial carotid occlusion, mTICI score, emergency carotid stenting, thrombectomy, onset-to-recanalisation time, arterial hypertension, HbA1c (%) and serum level of low-density lipoprotein cholesterol (mg/dl). Propensity score matching was carried out for all covariates that showed a statistically significant association with the grouping variable and the outcome on this regression model. The parameters are described in detail in *Supplementary Methods.* Propensity scores were created using logistic regression. One-to-one nearest neighbour matching with caliper adjustment was applied. The maximum allowed difference in propensity scores for matching (caliper value) was aimed at ≤0.2 (fraction of standard deviation of the logit of the propensity score) to ensure high quality matching. Visual and analytical comparisons were made to assess the quality of the matches. We calculated the standardised difference of covariate means and the distribution of covariates between groups to assess the overall balance of covariates between two groups. We aimed for a standardised difference of 10% (0±0.1) and a variance ratio of 1±0.2 to ensure a good balance. The average treatment effect was estimated.

In the sensitivity analysis, we performed multivariable lasso logistic regression to assess the association between serum HDL-C levels and imaging indices of reperfusion injury and favourable functional outcome adjusting for clinically relevant covariates. Residuals were tested for normality.

We carried out a shift analysis to compare the groups with low and high serum HDL-C levels. We used ordered multivariate logistic regression to assess the relationship between HDL-C and functional outcome as defined by the mRS score ranging from 0 to 6.

Available case analysis was carried out. Statistical significance was set at p<0.05. All analyses were performed using Stata® (StataCorp. 2021. Stata Statistical Software: Release 17. College Station, TX: StataCorp LLC).

### Ethical Standard

Our study was approved by the local institutional review board (Ethikkommission an der TU Dresden, IRB reference number: EK 272072017). Written informed consent for participation was waived in accordance with the national legislation and the institutional requirements.

## Results

### Study Population

Out of 1702 patients assessed for eligibility, we included 807 patients treated with thrombectomy for acLVO (420 females, median age 77 years [interquartile range, IQR 66-84]); median baseline NIHSS 15 [IQR 10-19]; median baseline mRS 5 [IQR 4-5]; median ASPECTS 7 [IQR 6-9], of whom 403 (49.7%) received preceding intravenous thrombolysis (IVT). Reperfusion injury was present in 192 (49.7%) of 386 patients with high serum HDL-C and in 123 (33.3%) of 369 patients with low serum HDL-C. The median mRS score at 90 days was 3 [IQR 1-4] in the high serum HDL-C group and 4 [IQR 2-6] in the low serum HDL-C group. The median NIHSS score at 90 days was 5 [IQR 1-14] in the high serum HDL-C group and 9 [IQR 2-19] in the low serum HDL-C group. Demographic and clinical characteristics, vascular risk profiles and imaging characteristics are detailed in Table 1. Subject selection and reasons for exclusion are shown in the study flowchart (Figure 1). The number of missing registry data was low. Details are given in *Supplementary Table S2*.

**Figure 1:**
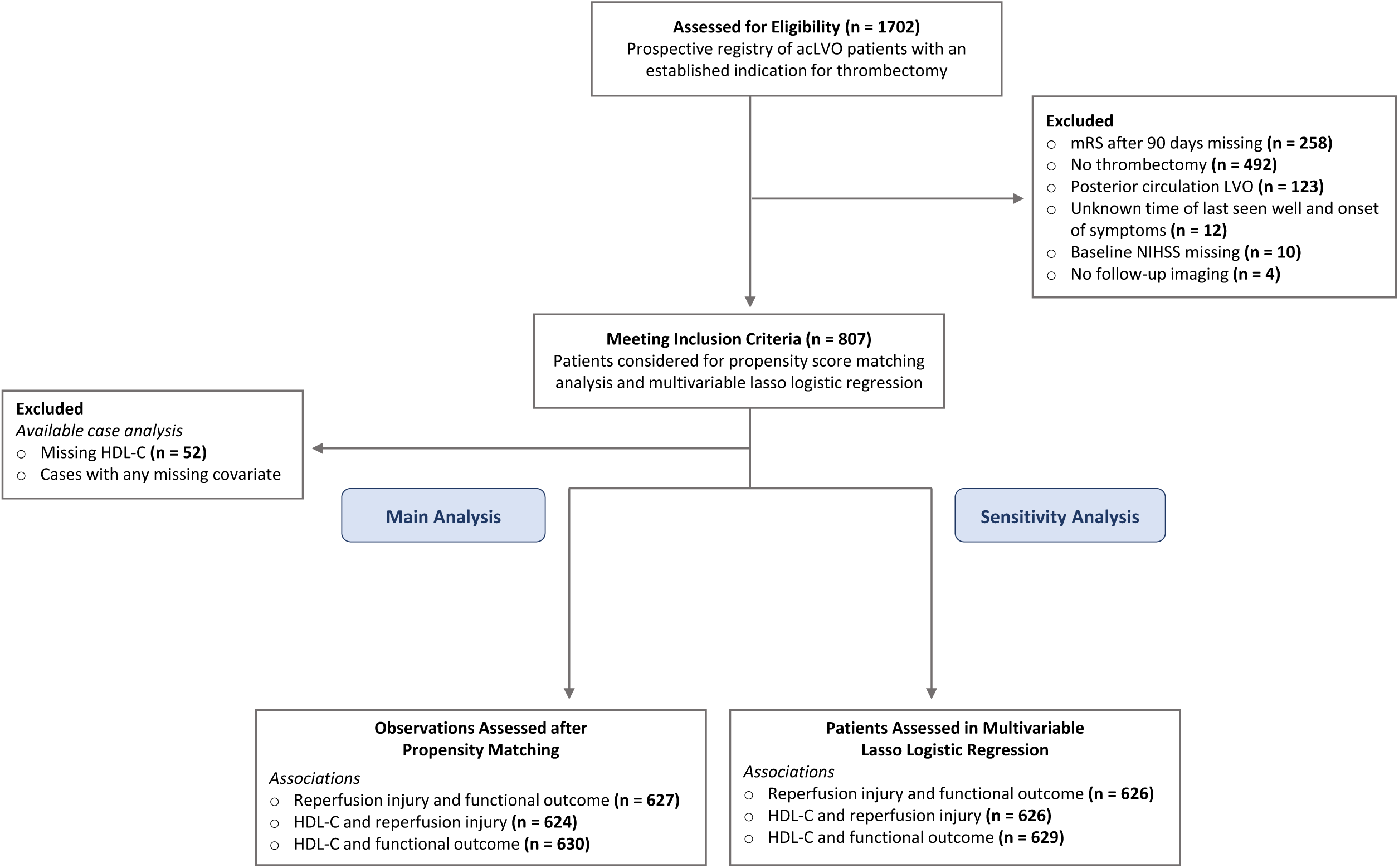
Study Flowchart. Study flowchart illustrating the screening and selection process of patients for inclusion in the main and sensitivity analyses of the study.

**Table 1:**
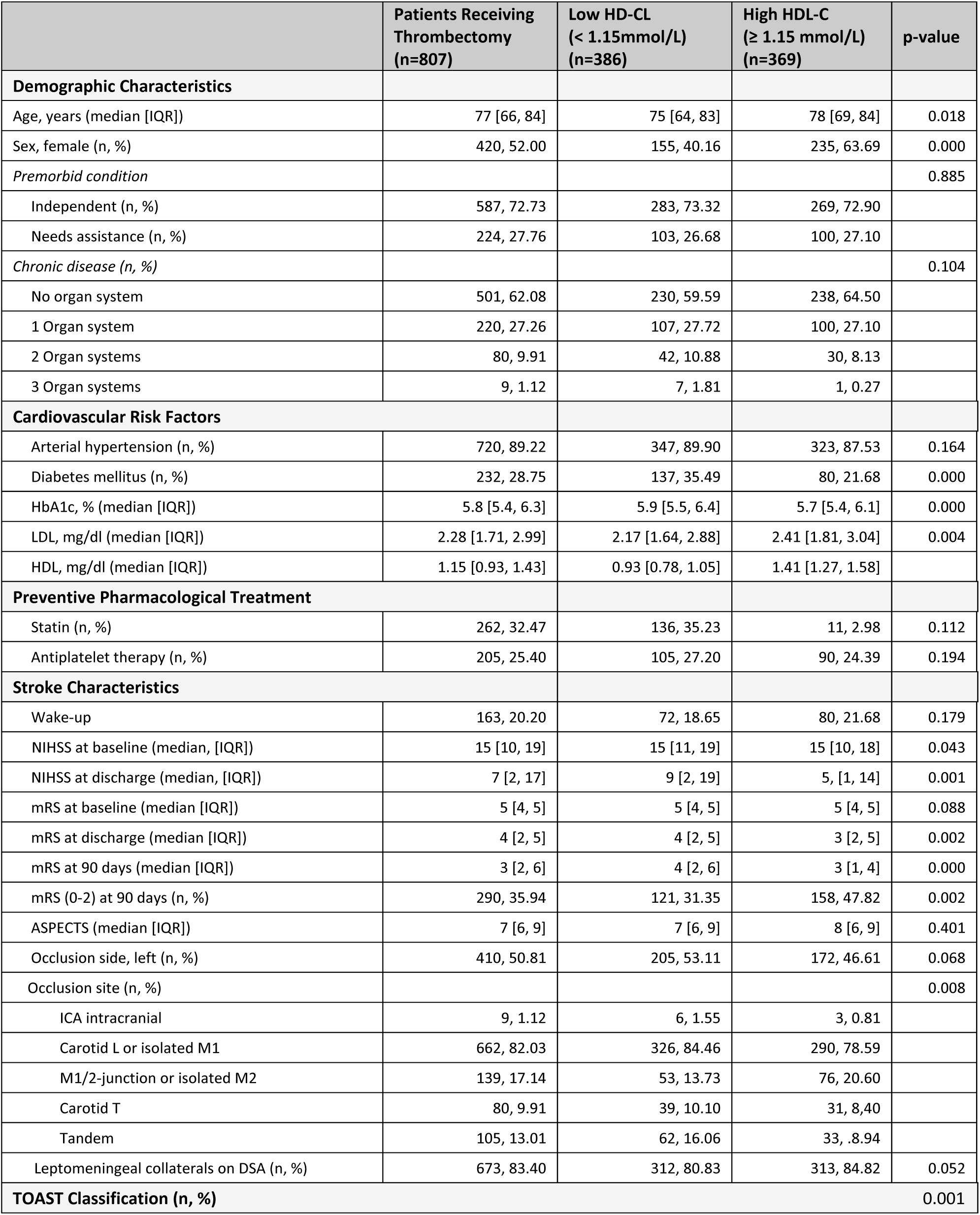
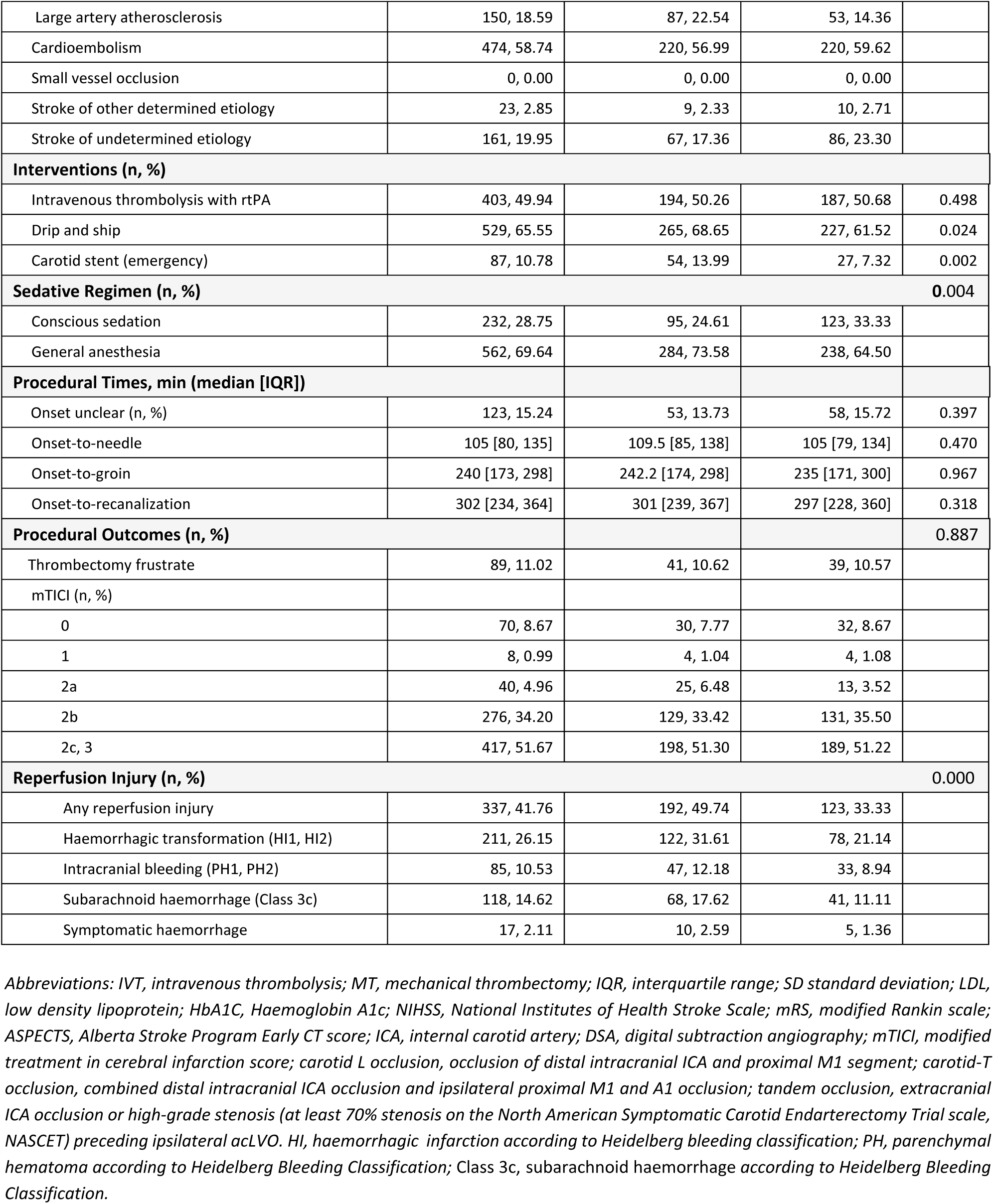
Demographic and Baseline Characteristics.

### Association of Reperfusion Injury with Functional Outcome

The presence of reperfusion injury on brain imaging was associated with a 14.8% increase in the probability of poor functional outcome (ß=0.15; 95CI% [0.06; 0.24]; *p*=0.001). The quality of the matching was good, as shown in *Figure 2A* and *Supplementary Table S3*. This observation was confirmed in the sensitivity analysis, where reperfusion injury was associated with higher odds of poor functional outcome (adjusted OR 2.69; 95% CI [1.75; 4.14]; *p*=0.000).

**Figure 2:**
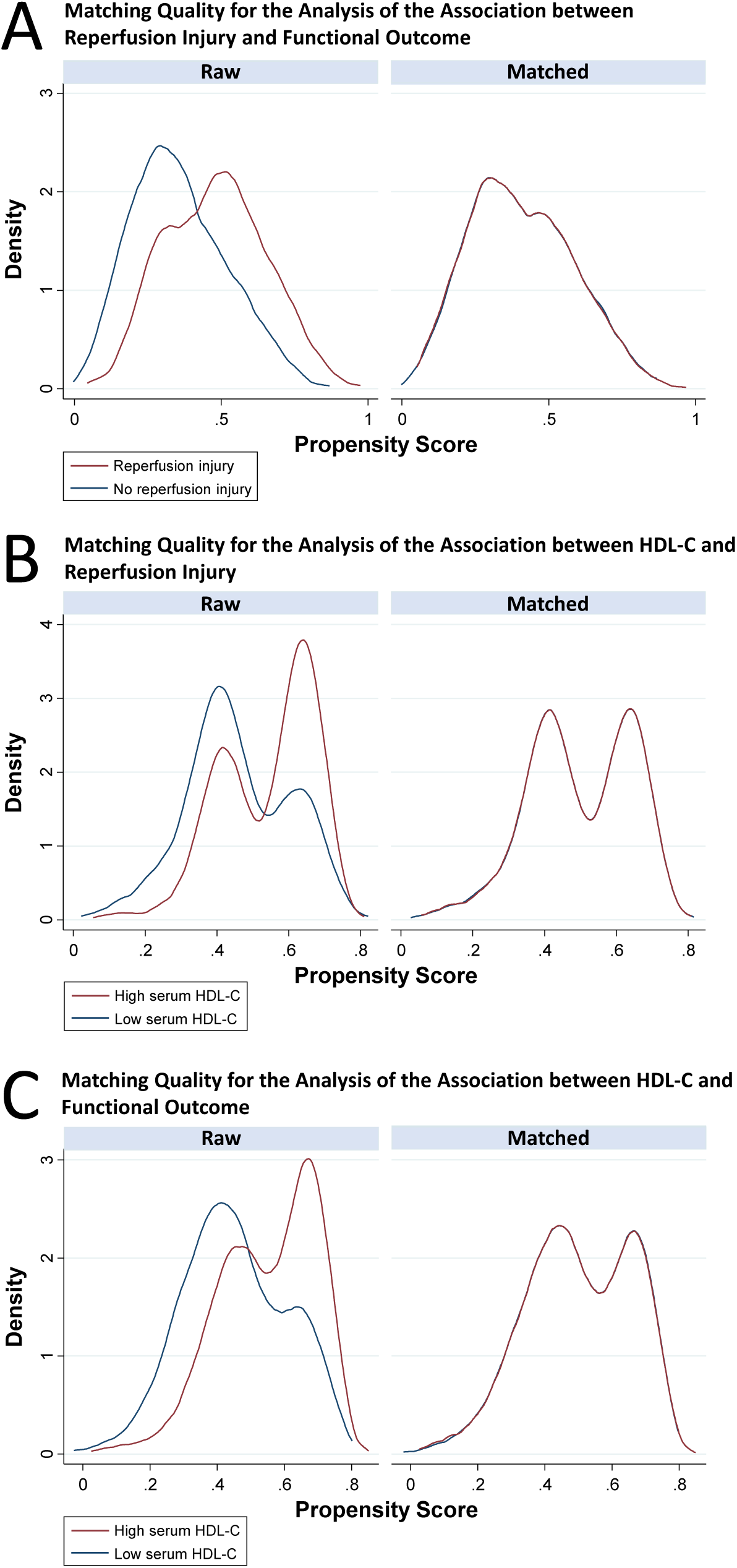
Matching Quality. Propensity score density plots for the analyses of A) the association between reperfusion injury and functional outcome; B) the association between HDL-C and reperfusion injury; and C) the association between HDL-C and functional outcome. The quality of matching was good in all cases.

### Association of Serum HDL-C Levels and Reperfusion Injury

A high serum HDL-C level was associated with a 13.6% reduction in the probability of reperfusion injury (ß=-0-14; 95CI% [-0.22; 0.05]; *p*=0.002). The quality of the matching was good, as shown in *Figure 2B* and *Supplementary Table S4*. This observation was confirmed in the sensitivity analysis, where a high serum HDL-C level was associated with lower odds of reperfusion injury (adjusted OR 0.62; 95% CI [0.43; 0.88]; *p*=0.008).

### Association of HDL-C Levels and Functional Outcome

A high serum HDL-C level was associated with a 13.2% increase in the probability of achieving a favourable functional (ß=-0.13; 95CI% [−0.22; −0.05]; *p*=0.003). The quality of the matching was good, as shown in *Figure 2C* and *Supplementary Table S5*. This observation was confirmed in sensitivity analysis, where a high serum HDL-C level was associated with lower odds of poor functional outcome (adjusted OR 0.60; 95% CI [0.40; 0.90]; *p*=0.002).

We observed a significant shift in the overall distribution of 90-day mRS scores in favor of the high HDL group over the low HDL group (adjusted OR 0.60; 95CI% [0.44; 0.81]; *p*=0.001), as shown in Figure 3.

**Figure 3:**
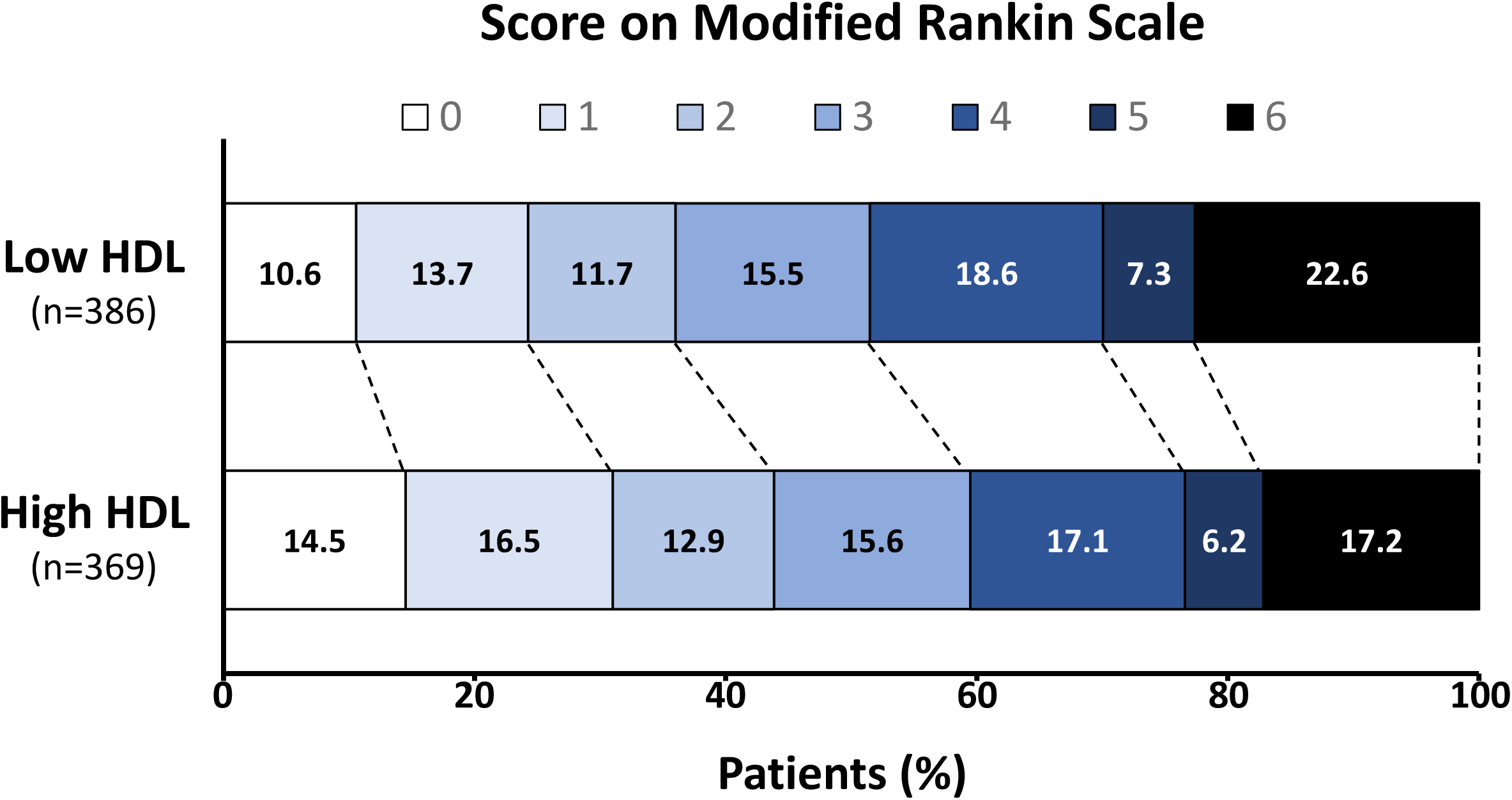
Shift Analysis. Shown are the scores on the modified Rankin scale at 90 days derived from ordered logistic regression with adjustment for age, sex, cardiovascular risk profiles, intravenous thrombolysis, baseline NIHSS, premorbid condition, chronic disease, onset-to-recanalization time, occlusion site, carotid stenting, modified treatment in cerebral infarction (mTICI) score, Alberta stroke programme early CT score (ASPECTS).

### Association of HDL-C Levels and Neurological Deficits at Discharge

A high serum HDL-C level was associated with a 2.3-point lower NIHSS score at discharge compared to a low HDL-C serum level (ß=-2.32; 95CI% [−4.14; −0.51]; *p*=0.01).

## Discussion

The main finding of this study is that in a prospective registry cohort of patients undergoing thrombectomy for acLVO, higher serum levels of HDL-C reduced the odds of haemorrhagic reperfusion injury and was associated with improved functional outcome at 90 days and a lower risk of early neurological deterioration. This observation suggests a protective effect of HDL-C at the time of the endovascular intervention.

The issue of reperfusion injury following thrombectomy has grown exponentially over the recent years due to the rapid expansion of the indications for this treatment. This is due to an extended time window from the onset of symptoms and a larger area of demarcated infarction that still allows the patient to be eligible for thrombectomy.^16–17^ As a result, the expansion of the indication has increased the possible amount of cumulative structural brain damage at the time point when reperfusion reaches ischaemic brain tissue. The potential clinical relevance of HDL-C-mediated protection against reperfusion injury is highlighted by a recent analysis of the prospective CIPPIS registry. This analysis showed that reperfusion injury was independently associated with poor functional outcome at 3 months in patients with successful recanalisation of acLVO^12^. We were able to reproduce this observation in our cohort, where patients with reperfusion injury after thrombectomy for acLVO had 2.87-fold higher odds of poor functional outcome.

Large randomised trials have evaluated the effects of candidate neuroprotectants such as nerinetide and uric acid on clinical outcomes in acLVO patients undergoing thrombectomy.^18–19^ Neither study provided sufficient evidence to legitimise the widespread clinical use of the investigational compounds tested. Recently, experimental strategies have been developed to specifically target and counteract reperfusion injury following thrombectomy. These approaches include ischaemic post-conditioning and pharmacological attenuation of NMDA receptor-mediated excitotoxicity and free radical toxicity. They are currently being tested in clinical trials.^20–21^ It is important to recognise that the pathways by which HDL-C may protect reperfused brain tissue from injury are complex and probably include preservation of the integrity of the blood brain barrier through activation of the scavenger receptor class B type I^11^, restricted overexpression of adhesion molecules by endothelial cells^10^, and attenuated neutrophil recruitment.^22^ However, these observations are based on animal studies. Their applicability to human brain tissue is unclear.

A recent study has translated insights from animal research on the cardioprotective effects of HDL-C into clinical research. The AEGIS-II trial investigated the effects of treatment with apolipoprotein A-1, which is the main protein in HDL-C, in patients with myocardial infarction and multivessel coronary artery disease. This randomised controlled study did not show superiority over placebo in preventing major adverse cardiovascular events.^23^ However, the mechanism of action of apolipoprotein A-1 in this indication is unlikely to translate to potential protective effects of HDL-C in the scenario of cerebral reperfusion injury. While cerebroprotective effects of HDL-C are likely to be mediated by protection of the blood-brain barrier through multiple mechanisms, the cardioprotective effects tested in AEGIS-II result from remodeling of intraluminal plaque formations in the coronary arteries by enhancing cholesterol influx via apolipoprotein A1. We focused our analysis on the haemorrhagic consequences of reperfusion injury in the brain and found consistent associations between higher HDL-C levels and reduced structural brain damage. Therefore, the potential protective effects of HDL-C may be strongest in preventing the final stages of cerebral reperfusion injury, when disruption of the blood-brain barrier leads to parenchymal haemorrhage.

Our study is subject to the limitations that are inherent to a non-randomized design. However, we used propensity score matching to balance covariates between the two groups created with high and low HDL-C serum levels and we were able to reproduce our observations on sensitivity analyses using multivariable regression models. We did not assess rates of expansion of the ischaemic core and penumbra over time to capture possible ischaemic manifestations of reperfusion injury. However, we focused our study on haemorrhagic complications of reperfusion injury in order to clearly capture a terminal manifestation of the pathophysiological cascade leading up to blood-brain barrier disruption. Hence, we cannot comment on the temporal dynamics of the development of the cerebroprotective effects of HDL-C in specific phases of this cascade. Our imaging outcome of reperfusion injury subsumes haemorrhagic transformation as well as parenchymal or subarachnoid haemorrhage. Parenchymal or subarachnoid haemorrhage apparent after thrombectomy may also result from intraprocedural complications of thrombectomy, i.e. vessel injury or perforation, potentially diluting the composite outcome. However, the prevalence of this interventional complication is low ranging from 0.6% to 5.5%^6,24^. It did not affect the robustness of the observations in our study. Reperfusion injury can manifest up to 72 hours after thrombectomy.^25^ We performed post-interventional follow-up CT within 24 hours and may have missed cases of late onset reperfusion injury. The generalizability of our findings may be limited by the monocentric design of our study. Lastly, the observed association between higher HDL-C levels and a favourable functional outcome at 90 days may be partly due to a long-term protective effect of HDL-C that is not related to the mechanisms of reperfusion. However, we also found an association between higher HDL-C levels and lower severity of neurological deficits at discharge, suggesting a predominance of an immediate effect related to reperfusion after thrombectomy.

## Conclusion

In patients undergoing thrombectomy for acLVO, higher serum levels of HDL-C reduced the odds of reperfusion injury, which translated into improved functional and neurological outcome. This association suggests that HDL-C has a previously unrecognised influence on the integrity of the blood-brain barrier during reperfusion in humans, which may influence the efficacy of thrombectomy and extend the role of HDL-C beyond traditional long-term cardiovascular protection.

## Data Availability

Data available on reasonable request.

## Acknowledgments, Sources of Funding, & Disclosures

## Acknowledgements

Acknowledgments

Daniel P. O. Kaiser is supported by the Joachim Herz Foundation. Timo Siepmann received grants from the German Federal Ministry of Health and Kurt Goldstein Institut, speaker fees from DACH-Gesellschaft Prävention von Herz-Kreislauf-Erkrankungen e.V., royalties from Astrazeneca for consulting and from Dresden International University for serving as a program director and a lecturer of the Master’s Program in Clinical Research. None of these activities were related to the current study.

## Supplementary Material

- Supplementary Methods
- Tables S1-S5
- References S1-S3
- STROBE checklist

## Non-Standard Abbreviations and Acronyms

AEGIS-II: ApoA-I Event Reducing in ischaemic Syndromes II
CIPPIS: Comparison Influence to Prognosis of CTP and MRP in AIS Patients
HDL-C: high-density lipoprotein cholesterol
IVT: Intravenous thrombolysis
AIS: acute ischaemic stroke
acLVO: anterior circulation large vessel occlusion
LDL-C cholesteorol: low-density lipoprotein
mTICI: modified Thrombolysis in Cerebral Infarction
STROBE: Strengthening the Reporting of Observational Studies in Epidemiology
NIHSS: National Institute of Health Stroke Scale
mRS: modified Rankin Scale
TOAST: Trial of Org 10172 in Acute Stroke Treatment
ASPECTS: Alberta Stroke Program Early CT Score
HI: haemorrhagic infarction
PH: parenchymatous hematoma.

